# Model-based impact evaluation of new tuberculosis vaccines in aging populations under different modeling scenarios: the case of China

**DOI:** 10.1101/2023.09.27.23296224

**Authors:** Mario Tovar, Joaquín Sanz, Yamir Moreno

## Abstract

The slow descent in TB burden, the COVID-19 pandemic, along with the rise of multidrugresistant strains of *Mycobacterium tuberculosis*, seriously threaten TB control and the goals of the End TB strategy. To fight back, several vaccine candidates are under development, with some of them undergoing the phases 2B and 3 of the development pipeline. The impact of these vaccines on the general population needs to be addressed using disease-transmission models, and, in a country like China, which last year ranked third in number of cases worldwide, and where the population is undergoing a fast process of demographic aging, the impact of TB vaccination campaigns may depend heavily upon the age of targeted populations and with the mechanistic descriptions of the TB vaccines. For these reasons, transmission models need to capture the coupling between TB dynamics and demographic evolution, as well as to be able to accommodate different mechanistic descriptions of TB vaccine protection. In this work, we studied the potential impact of a new TB vaccine in China targeting adolescents (15-19 y.o.) or elderly people (60-64 y.o.), according to varying vaccine descriptions that represent reasonable mechanisms of action leading to prevention of disease (PoD), or prevention of recurrence (PoR), each of them targetting specific routes to TB disease. To measure the influence of the description of the coupling between transmission dynamics and aging in TB transmission models, we explored two different approaches to comptute the evolution of the contact matrices, which relate to the spreading among different age strata. Our results show that the magnitude of model-based impact estimates substantially depends upon the vaccine profile, and it is also strongly related to the modeling approach chosen to describe the time evolution of contact matrices. In spite of these sources of uncertainty, our results also show, in line with previous modeling works, that elder vaccination is a suitable option in China to reduce the incidence of TB.

## I. INTRODUCTION

Tuberculosis (TB) is an infectious disease caused by the bacterium *Mycobacterium tuberculosis (M*.*tb*.) that usually affects the lungs. It is a complex but preventable disease with a high global burden that requires early detection and long treatments, and that is still among the leading causes of death for a single pathogen worldwide. In 2014, the World Health Assembly introduced the World Health Organization (WHO) “Global Strategy and Targets for tuberculosis prevention, care, and Control after 2015”, labeled as the End TB Strategy, which consists of completing a reduction of TB incidence and mortality rates by 90% and 95%, between 2015 and 2035 [1].

Arguably, the measures and interventions currently under use for TB control are effective, but also insufficient to meet the goal of the End TB strategy. Thus, although a decay in TB incidence and mortality has been achieved worldwide since 1990 [2], its yearly rate of reduction is still too slow. Furthermore, during 2020 and 2021, and for the first time in decades, the world witnessed a surge in global TB burden levels with respect to previous years due to the COVID-19 emergency that led to underdiagnosis and under-treatment of TB, along with the saturation of most healthcare systems [3–6]. During those years alone, the WHO estimated that TB was the cause of death of more than 1.5 and 1.6 million people respectively, worldwide, combining HIV-negative and positive cases [3, 7].

The recent increase in TB burden observed in 2020-2022 due to the irruption of the COVID-19 pandemic, threatens, in high-burden countries like India or Indonesia, to raise the TB-related mortality back to even higher levels than before in the next few years [8, 9]. Moreover, the ever-increasing rates of emergence of drug resistance [10] evidences the necessity of new tools against the disease, including new and better drugs as well as improved diagnosis methods. Among these new resources, the development of a new vaccine that either boosts or replaces the current bacillus Calmette-Guerin (BCG) is commonly referred to as the potentially most impactful single intervention to halt TB transmission, given the limited and variable efficacy levels observed for BCG against the more transmissible respiratory forms of the disease in young adults [11]. Consequently, the TB vaccine development pipeline is populated by a number of novel candidates of different types, based on a variety of immunological principles and vaccine platforms [12]. For estimating and comparing the potential impact of each of these candidates on halting the TB transmission chain, the development of epidemiological models arises as a powerful tool. Refining these models and addressing the main sources of uncertainty and bias in their architecture constitutes an important step toward the development of new TB vaccines.

In this work, we forecast the impact of the introduction of a new TB vaccine in China, which, as of 2021, represented 7.4% of the total number of TB cases worldwide [3]. To produce robust estimates for vaccine impact in this country, it is important to consider aging as a key demographic determinant where China differs from the majority of high-burden countries in TB. According to the UN population division estimates, among the top-8 countries with the highest number of incident TB cases in 2021 (China, India, Indonesia, Philippines, Pakistan, Nigeria, Bangladesh, and the Democratic Republic of Congo), China is the one where population aging in the years to come will be more pronounced, going from a median age of 39.0 in 2023 to 50.7 years in 2050. Considering this, recent modeling studies have suggested that targeting elder population groups in a vaccination campaign may produce a greater impact than targeting children or young adults [13].

Estimating the impact of a vaccine on an aging population must be done considering several technical aspects. From a modeling perspective, demographic aging couples with TB transmission dynamics critically in the age-mixing contact matrices [14]. These objects capture the relative frequency of social and/or physical contact between individuals of different age strata and constitute a great tool for representing contact patterns within epidemic models. When working with TB, whose time scales are comparable to those of demographic evolution, models need to incorporate sensible heuristics to describe the evolution of those matrices over time, which appears as a consequence of demographic evolution itself. In previous works [15], we identified different methods that can be used to adapt the contact matrices that were measured in a given population, as the population ages over time. Two of these methods are often found in modeling studies of TB [13, 14, 16]. First, contact matrices can be adapted to ensure that the symmetry of the encoded information is preserved, namely, the number of contacts per unit of time between two age groups, *i* and *j*, should be the same, when calculated from the number of contacts per capita, from *i* to *j* and from *j* to *i*. This method is commonly referred to as the pairwise correction method and ensures symmetry in reported numbers of contacts but does not produce contact networks whose mean connectivity is stable over time. To solve this issue, these matrices can be further corrected to ensure that not only the symmetry but also the average connectivity of the networks is preserved across time while underlying populations are aging. This second method, firstly proposed in [15], is referred to in this study as the intrinsic connectivity method, and produces contact structures that feature controlled contact densities on average, which are stable over time. The adoption of each of these methods to describe the evolution of contacts within TB transmission models may affect model outcomes regarding the impact of a new vaccine.

Another relevant aspect through which populations’ aging and vaccine impact forecasts may couple in ways that are hard to predict a priori is the vaccine mechanisms of action, combined with its protection profiles. A successful TB vaccine may confer either POD or POR through a variety of mechanisms, including halting fast progression towards primary TB upon a recent, first infection event, diminishing the endogenous reactivation rates upon latent infection, diminishing the risk of reinfection and/or preventing recurrence after recovery of a previous disease episode. Furthermore, the ability of a vaccine to confer protection through each of these mechanisms may depend upon the previous exposure of vaccine recipients to *Mycobacterium tuberculosis*, ascertained by an interferon-gamma release assay (IGRA). Since the fraction of individuals who have been previously exposed to the pathogen varies across age, and, in an aging population, that changes across time, exploring the effects of vaccine protection profiles and mechanisms of action at once is crucial to compare different vaccination strategies targeting different age groups in China.

The goal of this study is to investigate the role of these aspects on the vaccine impact foreseen from two vaccination campaigns targeting adolescents (15-19 y.o) and elder individuals (60-64 y.o.) in China, starting in 2025. Capitalizing on previous TB transmission models developed by the authors [14, 16], we produced in-silico impact evaluations of a series of vaccines with different protection profiles, acting through different mechanisms, evaluated in different simulations where contact patterns evolve according to different methods. In what concerns the distribution of vaccine-mediated protection across vaccinated individuals, we modeled vaccines as all-or-nothing (AON), as schematized in Figure 1, as it is widely used in the modeling literature [17–19]. AON vaccines confer perfect protection to a fraction of the vaccinated individuals but are ineffective for the remaining fraction of vaccinated individuals. The share between those fractions is related to the overall efficacy of the vaccine. To estimate the impact, we make use of two runs of the mathematical model (control and vaccine), and we analyze the results in terms of the targeted population, the protection profile of the vaccine, and the method used for updating the contact matrix.

**FIG. 1.**
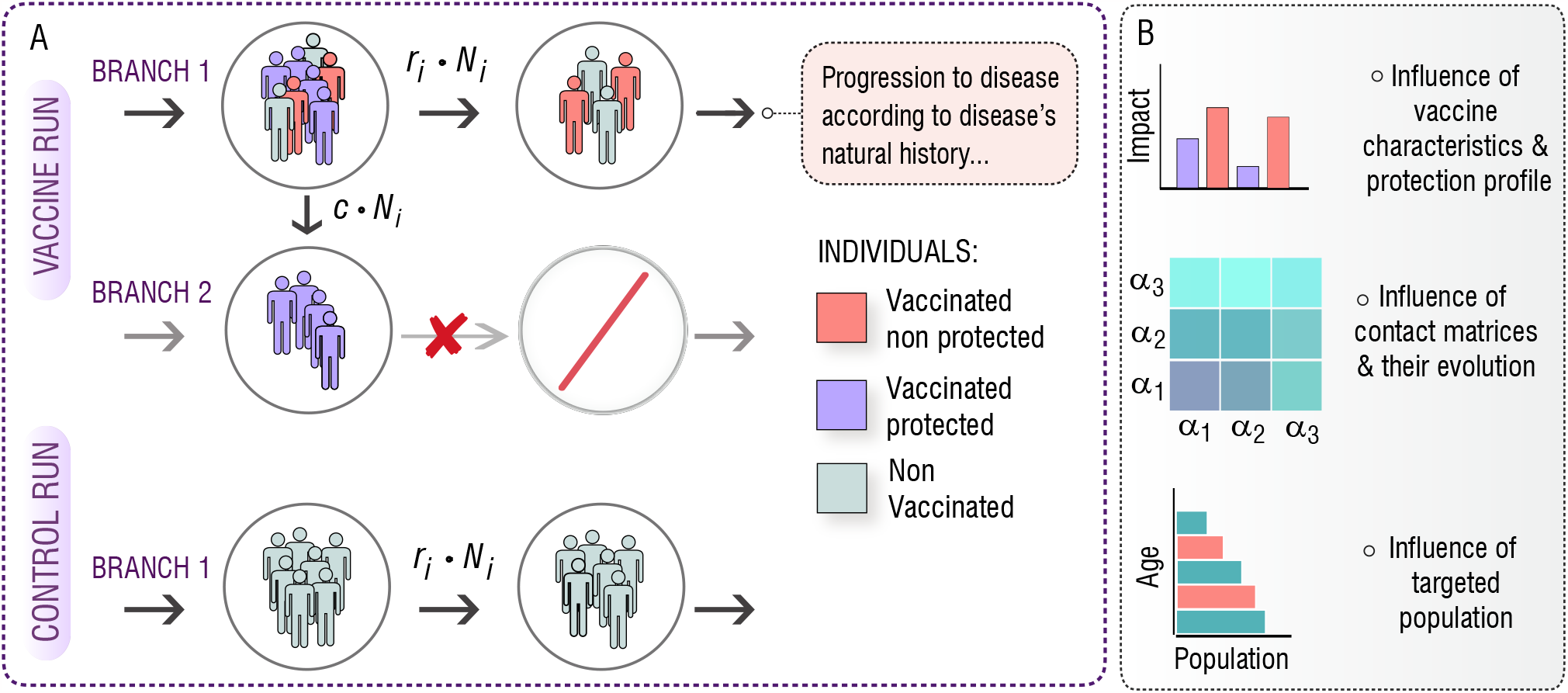
A. In-silico simulations for the introduction of an AON vaccine in a mathematical model of TB transmission. The model is run twice, one as a control run and the second time with the introduction of the vaccine. In the control run (bottom) the natural history of the disease remains unaltered, and every transition between states of the model is present. In the vaccine run, there are two different branches that evolve in parallel. On branch 1, the natural history of the disease remains unaltered, but a *c* fraction of the individuals that receive the vaccine is moved towards branch 2, in which the natural history is modified according to the effect of the vaccine, thus, neglecting some transitions and conferring protection. This way, a fraction, *c*, of the vaccinated individuals becomes protected. B. Implementing this general approach for different vaccine characteristics and protection profiles, under different descriptions of contact matrices evolution, we aim to study the effect of these aspects on the impact foreseen by our computational model for a vaccine applied either in adolescents (15-19 years old), or in older individuals (60-64 years old).

## II. MATERIALS AND METHODS

### A. Modeling the effect of the vaccine

The efficacy estimates obtained for a given vaccine in a clinical trial can be mapped onto mechanistic descriptions within a transmission model in a number of different ways. Elucidating what are the specific mechanisms at place that are most likely compatible with trials’ results for a given vaccine is not a trivial task, and the architecture of the model, as well as the characteristics of the population enrolled and detailed data analysis of the results are needed for extrapolating the efficacy levels observed in a trial into transmission models describing TB dynamics in entire populations[16, 20]. In a disease such as TB, a vaccine conferring POD or POR may base its protective effects on interfering with different processes throughout the natural history of the disease, preventing individuals’ progression to TB by halting specific routes to disease. In the lack of direct evidence concerning the specific dynamic mechanisms in place in a given vaccine, modelers often implement vaccine descriptions where all the main routes to disease putatively affected by the vaccine are equally impacted. Instead, in this work, we aim to compare the impact of different vaccines whose protection acts through different dynamical mechanisms. Formally, we assume that a vaccine can reduce the risk of progressing further from a given state toward disease, thus conferring protection to vaccinated individuals at different stages in the natural history of the disease. Capitalizing on our model, we identify four different basic vaccine mechanisms that can act either alone or combined:

- *E*_*p*_: Protection against primary TB: The vaccine confers protection against fast progression to disease upon a recent first infection event. This mechanism is present in a POD vaccine that prevents fast latency toward active disease (see supplementary appendix).
- *E*_*rl*_: Protection against endogenous reactivation of LTBI: Vaccine confers protection against endogenous reactivation of bacilli in individuals with latent TB infection (LTBI). This mechanism is present in a POD vaccine that prevents slow latency towards active disease.
- *E*_*q*_: Protection against TB upon reinfection: Vaccine confers protection against exogenous reactivation caused by a secondary infection event in subjects who had been previously infected. This mechanism is present in a POD vaccine that prevents progression towards active disease upon reinfection, for individuals who had already been exposed to the pathogen before (either LTBI or recovered individuals).
- *E*_relapse_: Protection against TB relapse: This mechanism is present in a POR vaccine that prevents endogenous reactivation in individuals who had a past episode of active TB.
- All: Every vaccine’s mechanism acts at the same time.

The interaction between vaccines conferring protection at any of the previous mechanisms, and the natural history of the disease is sketched in Supplementary Figure 3. In short, the fraction of the vaccinated individuals who get protection from the vaccine will face a modified version of the natural history according to the effect of the vaccine, where some key transitions are halted.

### B. Model-based impact evaluations of TB vaccines

We estimate the impacts of vaccines using an adapted version of the model in [14], which is a deterministic, age-structured model based on ordinary differential equations, where individuals belonging to different age strata are considered to experiment different levels of epidemiological risk. This translates, in general, into age-specific parameter values, as described in the supplementary appendix. The architecture that defines the disease dynamics within each age group represents the natural history of TB (see Supplementary Figure 1). The model also includes aging dynamics, which is key in countries undergoing fast demographic changes (see Supplementary Appendix, and Supplementary Figure 2).

Regarding the impact evaluation, we use two different runs of the model that ultimately lead to an estimate of the incidence rate reduction due to the vaccine. In the first run, specific values of all the epidemiological parameters are stochastically drawn from suitable distributions. Using the specific set of parameters obtained, the model is calibrated and the spreading of the disease is forecasted in a non-intervention scenario, referred here as the control run. Then, the model is run again, using the same calibration, but introducing the vaccine in 2025. This vaccine run does not follow qualitatively the same Natural History as in the control run, as the vaccine alters it by reducing the progression risk of protected individuals in certain transitions that depend on the characteristics of the vaccine (see supplementary appendix). Finally, the impact of the vaccine is estimated by comparing those two runs through the obtention of the incidence rate reduction (IRR) at the end of 2050, as follows:

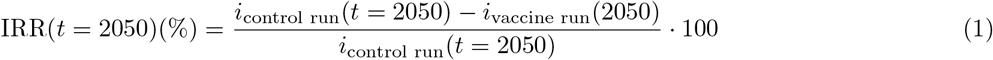

Repeating this procedure a number of *N* = 500 times, we obtain a distribution of forecasted vaccine impacts, which allows us to build suitable expected values and confidence intervals that propagate uncertainty from model inputs to vaccine impacts.

As already mentioned, all vaccines are introduced into the model according to an all-or-nothing scheme, which means they only show efficacy in a fraction *c* of the vaccinated individuals, which in this context represents the vaccine efficacy under a scenario of perfect coverage. The remaining 1*−c* fraction of vaccinated individuals do not benefit from these effects and preserve the same dynamics as the unvaccinated individuals. Formally, this is modeled by displacing a fraction *c* of vaccinated individuals from a control branch to a vaccine-protection branch, where the dynamics is modified to reflect these changes.

The vaccines considered in this work feature different levels of waning. As vaccinated individuals age from their age at vaccination *a*_*v*_ to *a > a*_*v*_, the vaccine efficacy is expected to decay, eventually becoming inefficient *w* years after vaccination. To implement vaccine waning in an all-or-nothing model, we introduced in the model a series of return fluxes that move individuals in the age group *a > a*_*v*_ back from the protected to the non-protected branch of the model. The intensity of those fluxes is given by Equation 2.

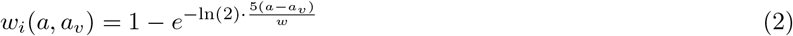

where *a* is the age group that suffers the waning, *a*_*v*_ is the age group that is being vaccinated, and *w* captures the waning, in years. This formula ensures that, after *w* years, the vaccinated individuals will suffer a waning intensity of 50%. Moreover, for any *a≤a*_*v*_, the waning intensity is set to zero, constituting a viable approach to implement vaccination campaigns targetting one specific age stratum. Then, the waning flux is calculated as:

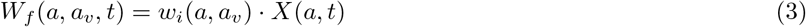

where *X*(*a*) is the population in the age group *a*, in the reservoir *X* at time *t*, where *X* is every reservoir that vaccinated individuals may lie into.

Besides the alterations in epidemiological risks experienced by the individuals protected by the vaccine, another important vaccine characteristic concerns the immunological status that individuals may need to fulfill before vaccination for the vaccine to confer protection to them. Depending on the immunological principles of the vaccine considered, its protection may unfold contingent on the previous exposure of the vaccinated subject to the pathogen, and, as such, it is possible that new vaccines may confer protection only to susceptible, immunologically naive individuals, or to individuals who have been previously infected with *M*.*tuberculosis*. To illustrate the effect of the dependency between the recipient’s status and vaccine efficacy, we reproduce vaccine impact simulations where the protective effect of the vaccine, described through the displacement of a fraction *c* of individuals towards the protected branch, only takes place from different source reservoirs. This way, we explore four possible scenarios where only a fraction *c* of individuals in the following reservoirs get protection: i) only susceptible individuals ii) only LTBI individuals iii) only previously exposed individuals (including LTBI and recovered subjects), and iv) all individuals.

Finally, the vaccination is implemented in two steps. First, a mass vaccination campaign, similar to the one proposed in[13] takes place, vaccinating annually a third of the population in the reservoirs affected by the vaccine, and for the age targeted population (15-19 or 60-64). Then, after this campaign vanishes, the vaccination continues routinely coupled to the aging.

### C. Updating contact matrices with evolving demography

Age-mixing contact matrices play a key role in epidemic spreading [21–23], as the complete knowledge of the network of contacts is usually unreachable or impossible to implement. Thus, for modeling purposes, it is useful to study agegroup interactions, where contact matrices indicate how age-strata mix between them. Usually, empirical contact matrices are obtained through statistical surveys. In these studies, participants are asked how many contacts they have during the day and with whom. This allows us to obtain the (average) number of contacts that an individual of a particular age *i* has with individuals of age group *j*. The resulting matrix is not symmetric due to the different number of individuals in each age group. However, it is precisely the demographic structure that imposes constraints in the entries of this matrix, as reciprocity of contacts should be fulfilled at any time (i.e., the total number of contacts reported by age-group *i* with age-group *j* should be equal in the opposite direction). Therefore, an empirical contact matrix, that has been measured on a specific population, should not be used directly without adapting it to the demographic structure of a different population under study.

This issue has important consequences in the field of disease modeling. As contact matrices play a key role in disease forecast, it is essential to assure that the matrices implemented are adapted to the demographic structure of the population considered to avoid biased estimations. For some short-cycle diseases like influenza, the time scale of the epidemic is much shorter than the typical times needed for a demographic structure to evolve [24]. The previous considerations are more troublesome for the case of persistent diseases that need long-term simulations, for which the hypothesis of constant demographic structures does not hold anymore [14]. Particularly, in the case of TB modeling, time scales are typically long, as the presence of latent individuals may lead to TB cases decades after primary infection [25]. This ultimately leads to the urge to adapt the contact matrices measured in a specific demography in such a way that they evolve accordingly to the demography of that setting. To this end, we capitalize on the methods proposed by Arregui et al. [15], which are briefly described below. We selected only methods labelled in the original article as M1 (Pairwise corrections) and M3 (Intrinsic connectivity) as the first one is typically used in the literature, also for modeling TB e.g. [13]), and is the simplest one for short-lives diseases, whereas the second one (M3) allows projecting contact matrices along with demography, which fits our needs in TB forecasting.

#### 1. Pairwise correction

The magnitude usually reported when measuring contact patterns is the mean number of contacts that an individual in age group *i* has with individuals in age group *j* during a measured period of time. Calling *M*_*i,j*_ this quantity, we observe that, in order to fulfill reciprocity, *M*_*i,j*_ should equal *M*_*j,i*_, which is not the case with directly measured data. An immediate correction is to average those numbers, so that the excess of contacts measured in one direction is transferred to the reciprocal. Then, the matrix entry in a new demography is computed as:

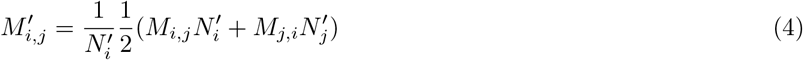

where 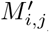 corresponds to the new demography under study. An example of the evolution of the contact matrix used in this study, under the pairwise correction is included in Supplementary Figure 4, panels A and B.

#### 2. Intrinsic connectivity matrix

An alternative method that preserves the mean connectivity of the contact network makes use of the density matrix or intrinsic connectivity matrix. Using the original data the density matrix Γ is extracted as:

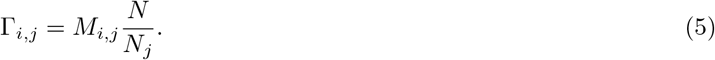

The Γ matrix corresponds, except for a global factor, to the contact pattern in a “rectangular” demography (a population structure where all age groups have the same density). Then, introducing a new demography, the contact matrix is obtained as:

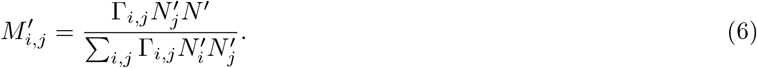

An example of the evolution of the contact matrix used in this work when using the intrinsic connectivity method is included in Supplementary Figure 4, panels C and D.

## III. RESULTS

After we implemented our computational model of TB transmission, we evaluated the impact of a TB vaccine of varying characteristics, simulated using models where contact matrices are updated using different methods, introduced in China in either adolescent (15-19 years old), or elder individuals (60-64 years old). In Figure 2 and Figure 3 we represent the forecast impact of each vaccine using our model. As discussed in the methods section, all vaccines are modeled according to an all-or-nothing scheme, conferring different types of protection (*E*_*p*_, *E*_*q*_, *E*_*rl*_, *E*_*relapse*_, or all at once, see Methods) to a fraction *c* of vaccinated individuals in certain disease reservoirs (susceptible, latent and/or recovered individuals, or all). The efficacy of the vaccine in all scenarios is set to *c* = 56%, as a reference value compatible with applying a highly protective vaccine with a 70% efficacy through a high-coverage campaign reaching 80% of the target population, similar to one among the most optimistic scenarios explored in previous modeling studies undertaken in China[13]. The vaccination campaign in the simulation starts in 2025, and we forecast the impact of the vaccine measuring the IRR (see methods) in 2050. Individuals of the targeted age group, are vaccinated when they first enter the corresponding age group. Furthermore, selected vaccines experience waning levels of 10 years, as described in more detail in the Methods section.

**FIG. 2.**
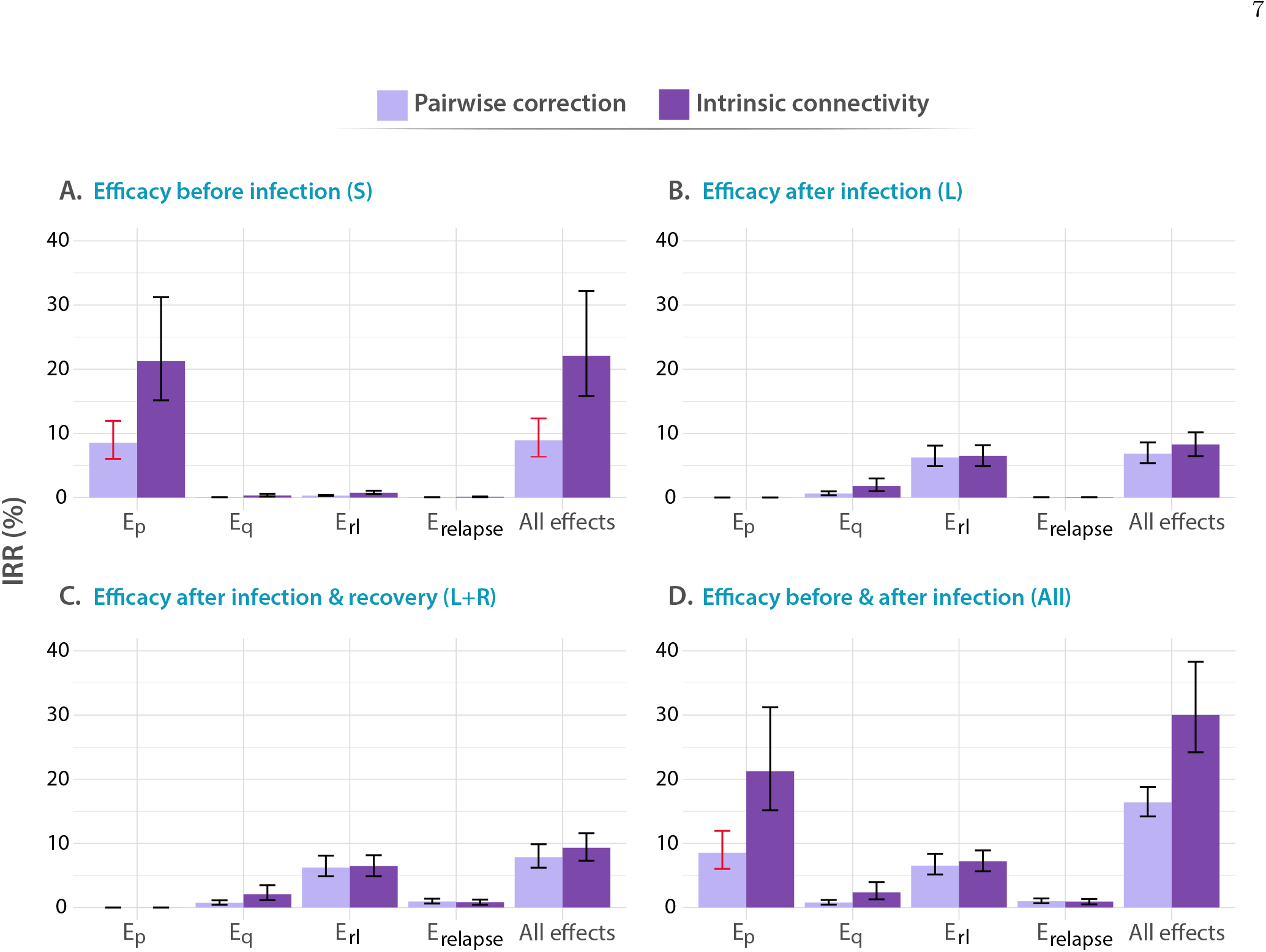
Impact of different vaccines applied to individuals between 60-64 years old. In all panels, tested vaccines act in different parts of the natural history of TB, halting progression to disease in one or more of the possible routes to disease, as described in the methods: *E*_*p*_: protection against primary TB, *E*_*q*_ protection against reinfection, *Er*_*l*_ : protection against endogenous progression to TB after LTBI, *E*_*relapse*_: prevention of recurrence. We analyze independently the impact of vaccines whose protective effects unfold when applied to individuals belonging to different compartments of the natural history, A. Susceptible subjects (efficacy observed before infection). B. Latently infected individuals (efficacy observed after infection). C. Latently infected and recovered individuals. D. Entire population. In each case, the impact of each vaccine is evaluated for a waning level of 10 years. In all panels, bars represent median values for the IRR measured in 2050, associated with the introduction of the vaccine in 2025. Error bars capture 95% confidence intervals from a set of *N* = 500 model outcomes in each case.

**FIG. 3.**
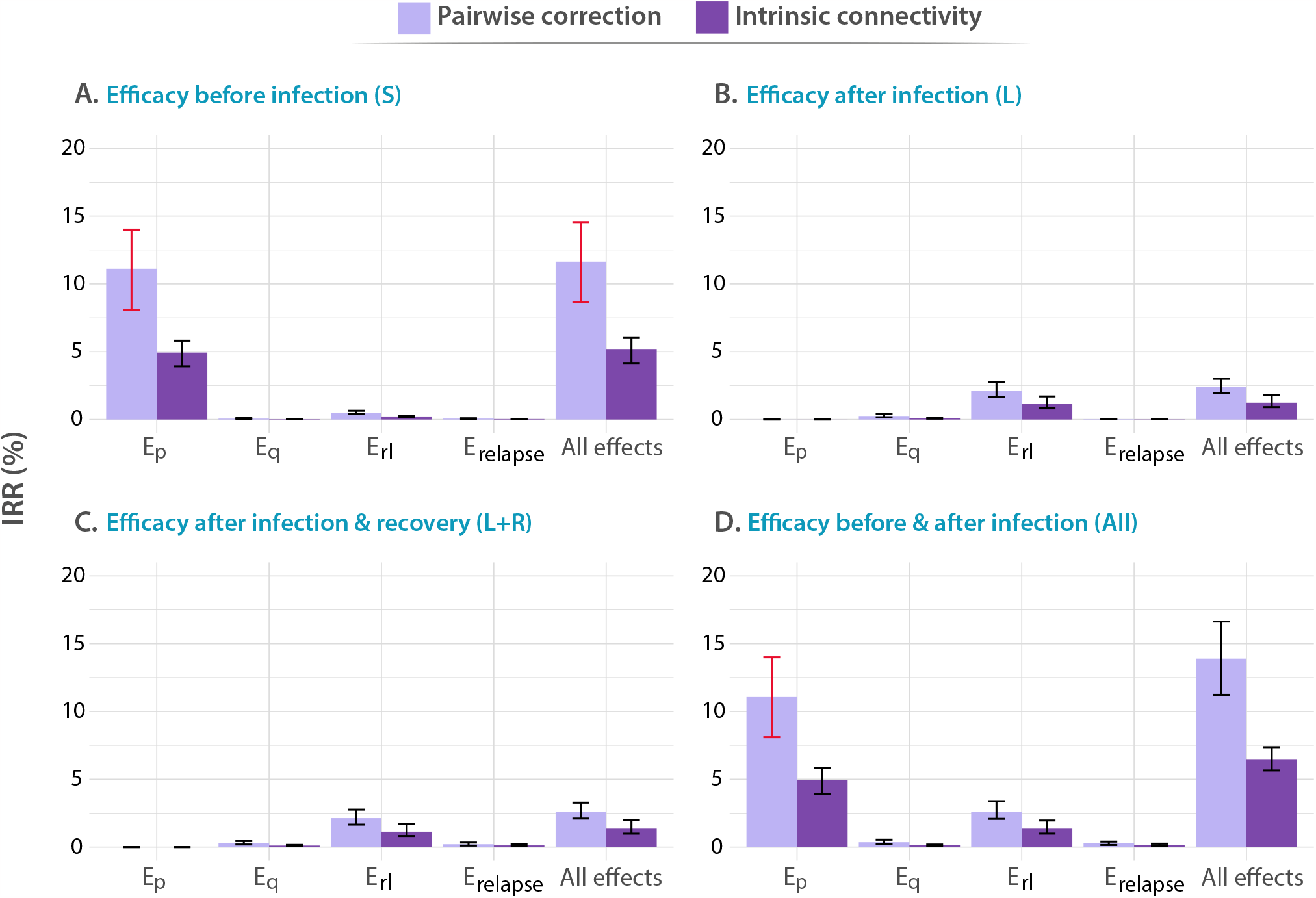
Impact of different vaccines applied to adolescents, with ages between 15-19 years old. Analogous to Figure 2, in all panels, tested vaccines act in different parts of the natural history of TB, halting progression to disease in one or more of the possible routes to disease: *E*_*p*_: protection against primary TB, *E*_*q*_ protection against reinfection, *E*_*rl*_ : protection against endogenous progression to TB after LTBI, *E*_*relapse*_: prevention of recurrence. We analyze independently the impact of vaccines whose protective effects unfold when applied to individuals belonging to different compartments of the natural history, A. Susceptible subjects (efficacy observed before infection). B. Latently infected individuals (efficacy observed after infection). C. Latently infected and recovered individuals. D. Entire population. In each case, the impact of each vaccine is evaluated for a waning level of 10 years. In all panels, bars represent median values for the IRR measured in 2050, associated with the introduction of the vaccine in 2025. Error bars capture 95% confidence intervals from a set of *N* = 500 model outcomes in each case.

In Figure 2, we gather the IRRs achieved by the different vaccines described above when applied to the elderly population. In this case, we found that, when vaccines are able to confer protection to immunologically naive individuals, either alone (Susceptible only) or along with the rest of individuals in the population, (whole population), vaccines featuring the largest impact are those that are able to prevent fast progression to primary TB upon recent infection (*E*_*p*_ vaccine featuring 24.9% IRR, 95% CI [17.8-36.1]) in panel A). Instead, if vaccine protection only unfolds on individuals who had been previously infected by the time of vaccination (protection active to either latent or latent plus recovered individuals only), the most impactful vaccines are those that are able to protect individuals from developing active TB upon endogenous reactivation of dormant bacilli (*E*_*rl*_ vaccine featuring 6.47% IRR, 95% CI[4.88-8.15] in panel C). Furthermore, it is important to notice that, in all vaccines tested leading to an impact higher than 1% under at least one of the two methods explored for describing contact matrices evolution, the impact was systematically higher when updating the contact matrix according to the intrinsic connectivity approach.

In turn, in Figure 3, we present the analogous results associated with a vaccination campaign targeting the population between 15 and 19 years old. Although vaccines protecting against primary TB are still more impactful as long as susceptible individuals are protected (*E*_*p*_ vaccine featuring 11.11% IRR, 95% CI [8.10-13.98]) in panel A, vs. other vaccines), and vaccines halting endogenous reactivation of latent bacilli are more impactful if protection takes place after infection, in this case, the impact associated with vaccines in the latter case is comparatively lower than what is found in elders (*E*_*rl*_ vaccine in panel C yields 2.14 IRR, 95% CI [1.65 2.76], when in elders yielded 6.47% IRR, 95% CI[4.88-8.15]). In what concerns the influence of contact matrices on forecast impacts, interestingly, we observe that the highest impacts were associated with the pairwise-corrections method, unlike what is observed in the elderly age group.

Admittedly, comparing the impacts from both campaigns targeting elders (Figure 2) and adolescents (Figure 3), we see that the question of what is the optimal age group to target in an immunization campaign for a new TB vaccine finds different answers depending on the combination of vaccine characteristics, protection profiles, and modeling assumptions. More specifically, in cases where previous infection is required for vaccines to elicit their protective effects, via protection against endogenous reactivation (*E*_*rl*_) or against relapse (*E*_*relapse*_), then targeting the older age group always appears as a superior choice according to our simulations (6.47% IRR, 95% CI[4.88-8.15] in most impactful vaccine in elders vs. 2.14 IRR, 95% CI [1.65 2.76] in adolescents). However, as soon as protection against primary TB is granted to susceptible individuals as one of the possible protective mechanisms of the vaccines, the quantitative description of contagion dynamics implemented within our model becomes more crucial, and, consequently, model forecasts are more sensitive to the adoption of either one of the two modeling approaches explored for describing contact matrices evolution: pairwise corrections vs. intrinsic connectivity. As a result, only in some occasions when the over-simplified pairwise correction method is adopted, the impacts foreseen for an adolescent-focused campaign can overcome the impacts found for an elder vaccination campaign (Bars marked in red) for these vaccines. For instance, an *E*_*p*_ vaccine yields 11.11% IRR (95% CI 8.10-13.98) under pairwise correction in adolescent, surpassing the 8.54% IRR (95% CI 6.03-11.95) obtained in elders for the same method, when vaccine protection unfolds before infection, whereas for the rest of vaccines, elders score higher impacts no matter what method is used.

In order to understand the influence of the vaccine mechanisms on their respective impacts, it is sufficient to analyze the time evolution of the distribution of TB cases across the different routes to disease classically described in TB, aggregated across age groups, as we represent in Figures 4A and 4B. These routes include fast progression to primary TB upon a first, recent infection event; TB after endogenous reactivation from LTBI; TB upon exogenous reinfection and, last, TB recurrence after a previous disease event (see supplementary methods). Importantly, each of the four vaccine mechanisms explored in this work tackles specifically each one of these routes. As seen in Figures 4A and 4B, primary TB upon recent infection is the prominent cause of TB cases during the simulated period, which makes protection against primary TB the most impactful vaccine mechanism, at least, as long as the susceptible individuals (who are those under a higher risk of developing primary TB upon infection[26]), could be protected by the vaccine (see Figures 2 and 3, panels A,D). Furthermore, we also observe that endogenous reactivation of LTBI individuals is the second type of event responsible for the highest share of TB cases, which in turn explains why vaccines protecting LTBI individuals are most impactful when they protect against endogenous reactivation (see Figures 2 and 3, panels B,C), and why vaccines targeting re-infection or relapse are comparatively less impactful, even when applied on older age groups where prevalence of infected and recovered subjects is higher.

**FIG. 4.**
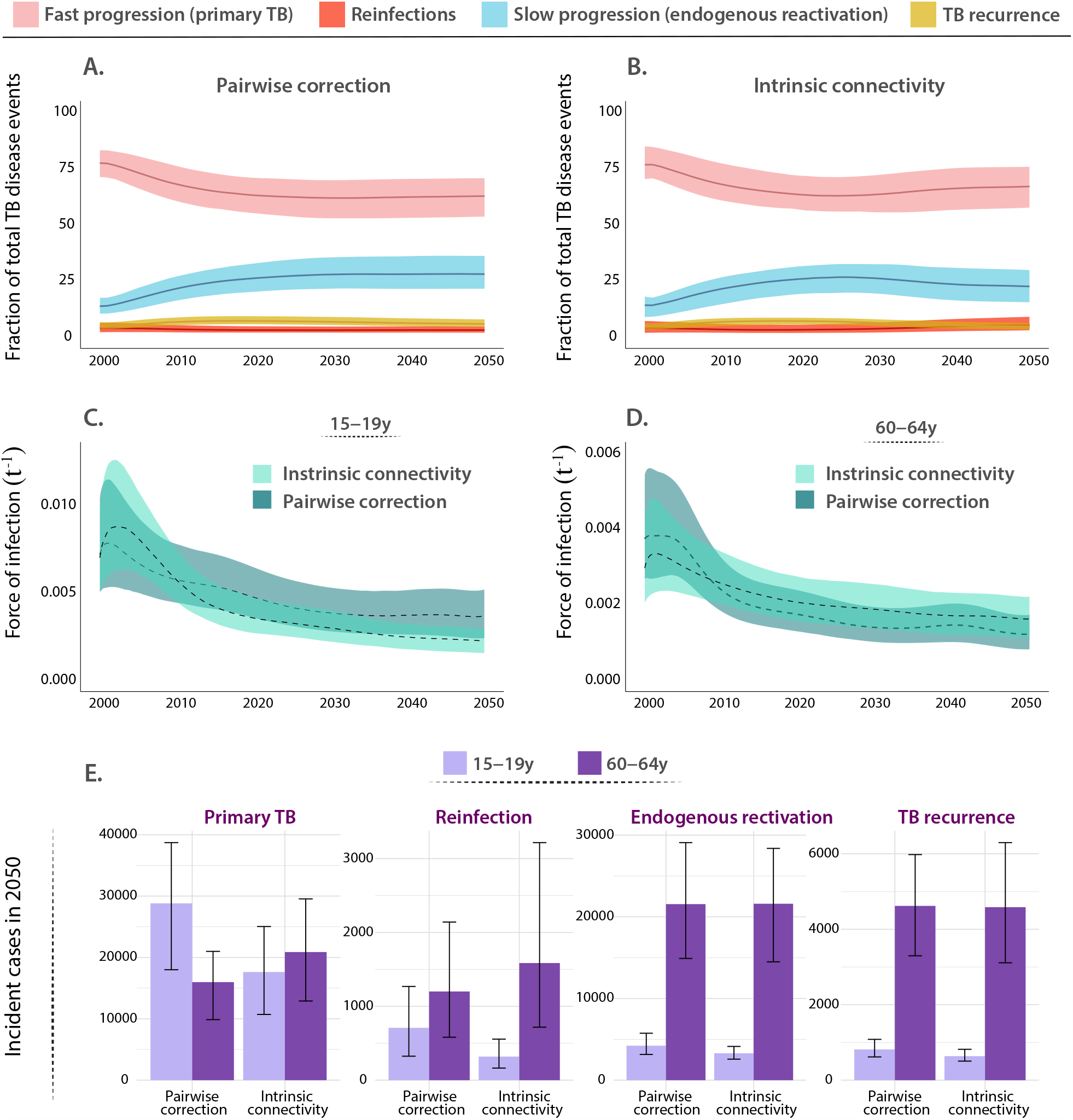
A,B. Evolution of the Percent of TB cases associated with rapid progression upon recent infection (primary TB), endogenous reactivation of LTBI, TB upon reinfection, or TB relapse; each of them foreseen from the indicated method for describing contact matrices evolution (pairwise correction or intrinsic connectivity). Primary TB upon recent infection, followed by endogenous reactivation of LTBI individuals are the two most common types of events. Central lines are medians and the shadowed areas represent the 95% confidence intervals from *N* = 500 model realizations. C,D. Evolution of the force of infection associated with individuals in age groups 15-19 and 60-64 in the period 2000-2050, as foreseen by the model when using the two frameworks for describing the time evolution of contact matrices. Adopting the simpler pairwise correction method results in a lower estimate of the force of infection in the older age group, as well as an overestimation in the younger stratum. Central lines are medians and the shadowed areas represent the 95% confidence intervals from *N* = 500 model realizations. E. Break down of the different contributions to the overall TB incidence pool in 2050, distributed across the routes to disease protected by the vaccines under study. The number of cases in the routes associated with already exposed individuals is systematically higher in elders than adolescents, no matter which correction for the contact matrices is at play. Bars represent median values measured in 2050. Error bars capture 95% confidence intervals from *N* = 500 model realizations.

Furthermore, in order to understand the role of the modeling assumptions concerning contacts when comparing impact forecasts from analogous immunization campaigns, it is important to highlight that their influence manifests more strongly when comparing forecasts for vaccines targeting TB upon recent infection (*E*_*p*_ vaccines) or reinfection (*E*_*q*_ vaccines), which is to say when the vaccine targets transmission. In such cases, the adoption of the most adequate method providing intrinsic connectivity control appears systematically associated with larger impacts when we analyze elder-focused campaigns, as well as lower impacts when we focus on campaigns targetting adolescents. This can be further contextualized by observing the evolution of the force of infection (defined as the fraction of susceptible individuals in a given group that gets infected per year) that individuals in each of these two age strata are subject to, according to each of the two models explored to describe the evolution of contact matrices. As seen in Figures 4C and 4D, using the pairwise model appears associated with an underestimation of the force of infection suffered by individuals in the older age group, and an overestimation of it among adolescents, with respect to the adoption of the more rigorous correction method based on preserving the intrinsic connectivity of contact matrices.

Finally, in order to contextualize the differences in impact found between elder and adolescents-focused campaigns, in Figure 4E, we present a simultaneous breakdown of TB cases predicted by 2050, in each of the age groups, associated with each of the routes to disease, according to each of the contact matrices models. In this figure, we see how both modeling approaches for contact matrices concur in assigning a higher incidence for TB cases related to LTBI reactivation, reinfection, or relapses, which can be interpreted as the main cause why, in Figures 2 and 3, we observe that *E*_*rl*_, *E*_*q*_ and *E*_*relapse*_ vaccines are systematically more impactful in elder individuals than in adolescents. However, in Figure 4E, we can also observe how the number of cases associated with primary TB is either higher, or lower in adolescents than it is in the older age group, depending on whether we adopt the pairwise correction method, or the intrinsic connectivity method, respectively.

Importantly, none of these observations are affected by the level of vaccine waning: while Figures 2 and 3 capture impacts associated with vaccines whose protection lasts ten years, largely comparable results are obtained for longer lasting vaccines, as summarized in the supplementary appendix (see Supplementary Figure 5, corresponding to waning=20 years).

## IV. DISCUSSION

Mathematical disease-transmission models are a powerful tool for estimating the impact of new TB vaccines, which, if done properly, may be instrumental in comparing the potential of different vaccine candidates and immunization campaigns. This is true, especially in TB, where vaccine development must face two simultaneous hindrances. First, vaccine efficacy is harder to foresee before phases 2b/3 of the development pipeline than for other diseases, given the lack of reliable correlates of immune protection[27]. Second, the architectures of the clinical trials of vaccine efficacy that are being adopted to test novel TB vaccine candidates are highly diverse [16, 20], and the protection profiles of the tested vaccines may be equally diverse. Taken together, these issues claim the development of rigorous computational models to produce impact comparisons for different vaccines tested in trials of different characteristics and implemented through assorted immunization campaigns. These constitute extremely non-trivial tasks, which enhances the need to ensure that current TB models can handle them while minimizing bias and uncertainty.

In accomplishing this goal, an aspect that demands special attention is the description of the coupling between demographic aging and the evolution of TB epidemiology in a given population. This is especially true in a country such as China, where two simultaneous aspects concur, namely: an intense process of demographic aging -already ongoing, and expected to continue in the next few decades-, concomitant with a high burden of TB incidence and prevalence levels. While previous works pointed to the observation that immunization campaigns targeting older age groups (paradigmatically individuals above 60 years old) are expected to cause a stronger reduction in global TB incidence levels than campaigns targeting adolescents (16-20 years old) [13], the robustness of these results under different modeling scenarios, including different vaccine characteristics and modeling decisions concerning the evolution of contact matrices among different age-groups remained to be proven.

Capitalizing on a mathematical model previously developed by the team [14, 16, 20], in this work we reproduce the general observation that, in China, immunization campaigns targeting older individuals, in the age group between 60 and 64 years old, are associated with promising levels of reduction in the incidence rates expected by 2050, with varying forecast impacts depending on vaccine characteristics and modeling assumptions, especially if the vaccine is able to protect already exposed individuals. This observation can be interpreted in light of the demographic shift expected in the country, where older age strata are expected to accumulate a higher fraction of total TB cases in the years to come. However, by using our model, we were able to address, for the first time in this study, how this observation may depend, in turn, on vaccine characteristics (the combination of its mechanisms of action and protection profile), as well as on modeling assumptions (the description of contact matrices over time).

On the one hand, when modeling TB vaccines, it is important to acknowledge the multiplicity of possible mechanisms of action a vaccine may confer protection through [16, 20]. This aspect, in turn, must be considered simultaneously with the fact that the initial immunological profile of vaccinated individuals (i.e. their IGRA status) may in turn influence the ability of the vaccine to provide its protective effects. [13]. In this work, we describe how these two aspects are coupled, generating strong interactions between the vaccine mechanisms in place and the sub-population reservoirs that may gain protective effects upon vaccination. Specifically, we observed that vaccines protecting susceptible, immunologically naive individuals are more impactful when their mechanism of action is based upon the prevention of primary TB after infection. This result can be understood by observing that progression to primary TB upon recent infection represents not just the main epidemiological risk for susceptible individuals, but the most common route to disease in the whole population, as sketched in Figures 4A and 4B. Importantly, our simulations indicate that tackling primary TB is the most promising intervention, not only when the immunization campaign targets adolescents, but also when it targets older individuals, as long as vaccine protection unfolds for susceptible individuals at least. Furthermore, when a vaccine requires that vaccinated individuals have previously been infected, the most impactful vaccine mechanism of action is based on preventing endogenous reactivation of LTBI. This indicates that, for LTBI subjects, endogenous progression to TB represents the highest epidemiological risk, which is shown in Figures 4A and 4B, where endogenous reactivation in the second most common route to disease. These couplings between vaccine mechanisms and protection profiles should be carefully taken into account when testing and comparing vaccine candidates with different profiles and immunization strategies.

On the other hand, in a country such as China, it is key to produce model-based descriptions of TB dynamics that are robust under the scenario of fast demographic aging. Under these circumstances, the adoption of plausible description frameworks to describe the evolution of contact matrices is key. The reason for this is that these matrices capture the relative frequency of contacts that may lead to new infections among individuals of different ages, and these are bound to evolve with time in an aging population. While relatively naive descriptions of contact matrices based on symmetry preservation through pairwise corrections are enough when modeling infectious diseases during short periods of time, TB demands more sophisticated approaches that preserve not only the symmetry but also the overall connectivity of the entire contact networks [15]. The reason for this is that during the extended time windows that TB modeling requires, demographic structures are expected to vary significantly, and, with them, the frequencies of social contacts among age strata, and the entire connectivity, measured as the average number of contacts per individual, of the system.

In this work, we showed that more sophisticated modeling approaches based on imposing the preservation of the intrinsic connectivity of contact networks (instead of simpler methods based on pairwise corrections aiming only at preserving symmetry) are linked to higher vaccine impacts when immunization campaigns target transmission among elder individuals. In turn, for campaigns targeting transmission among adolescents, it is the simpler methods, based on pairwise corrections, the one yielding higher impacts. In short, our simulations indicate that vaccines whose protection mechanisms take place after infection (e.g. *E*_*rl*_, *E*_*q*_ and *E*_*relapse*_ on L, L+R, or All population), are expected to elicit higher population impacts if applied in elder individuals, as well as vaccines protecting susceptible individuals against primary TB, providing that an adequate modeling approach is used to describe the evolution of their contact matrices, ensuring intrinsic connectivity control.

We also need to mention that our approach is not exempt from limitations that affect TB transmission models. The outcomes of our model depend on a series of epidemiological parameters and initial burden estimates that are subject to strong sources of uncertainty, thus propagating this uncertainty to the results. This means that future improvements in measuring the input data are expected to impact the quantitative outcomes of our mathematical model, in the same way it would affect any other model that leans on them. Always bearing in mind the strong uncertainties that the forecasts inherit, our results highlight the importance of acknowledging the complexity of TB transmission dynamics when modeling the effects of an age-focused intervention such as the introduction of a new vaccine on a specific age group.

In closing, our results emphasize the idea that immunization campaigns for the introduction of new TB vaccines in different countries can be, and must be, tailored using mathematical models that integrate information on vaccines’ profiles, population demography, and basal TB epidemiology.

## CONFLICT OF INTEREST STATEMENT

The authors declare that the research was conducted in the absence of any commercial or financial relationships that could be construed as a potential conflict of interest.

## AUTHOR CONTRIBUTIONS

M.T, J.S and Y.M designed the study. M.T performed the analysis and simulations that support the findings. M.T, J.S and Y.M analyzed and discussed the results. M.T and Y.M. wrote the manuscript. All authors revised and approved the submission of the final version of the manuscript.

## FUNDING

We thank M. Clarin for help with the figures. M.T. acknowledges the support of the Government of Aragón through a PhD contract. J.S. acknowledges support from the Spanish Ministry of Science and Innovation (MICINN) through grant PID2019-106859GA-I00 and Ramón y Cajal research grant RYC-2017-23560, as well as to the Government of Aragón, Spain, and ‘ERDF A way of making Europe” through grant B49-23R (NeuroBioSys). Y.M was partially supported by the Government of Aragón, Spain, and “ERDF A way of making Europe” through grant E36-23R (FENOL), and by Ministerio de Ciencia e Innovación, Agencia Española de Investigación (MCIN/AEI/10.13039/501100011033) Grant No. PID2020-115800GB-I00. The funders had no role in study design, data collection, and analysis, decision to publish, or preparation of the manuscript.

## DATA AVAILABILITY STATEMENT

The data that support the findings of this study are publicly available at the original sources. The code that supports the findings of this study is available from the corresponding author upon reasonable request.

## Notes

### Competing Interest Statement

The authors have declared no competing interest.

### Funding Statement

M.T. acknowledges the support of the Government of Aragon through a PhD contract. J.S. acknowledges support from the Spanish Ministry of Science and Innovation (MICINN) through grant PID2019-106859GA-I00 and Ramon y Cajal research grant RYC-2017-23560, as well as to the Government of Aragon, Spain, and `ERDF A way of making Europe'' through grant B49-23R (NeuroBioSys). Y.M was partially supported by the Government of Aragon, Spain, and ``ERDF A way of making Europe'' through grant E36‐23R (FENOL), and by Ministerio de Ciencia e Innovacion, Agencia Espanola de Investigacion (MCIN/AEI/10.13039/501100011033) Grant No. PID2020‐115800GB‐I00.

