## Supplementary Material for "Model-based impact evaluation of new tuberculosis vaccines in aging populations under different modeling scenarios: the case of China"

(Dated: September 27, 2023)

### CONTENTS

Supplementary Methods 1

Supplementary results 6

Supplementary References 7

### SUPPLEMENTARY METHODS

For this study, we used a modified version of Arregui et al.'s model[1] to assess the long-term effects of vaccination in TB burden in China. Our version of the model retains the original's fundamental structure, but improves upon the method of calculating confidence intervals to better account for uncertainties, and allows to introduce a wide variety of vaccine descriptions. This is a deterministic, age-structured model that uses ordinary differential equations to differentiate between levels of epidemiological risk within age groups. While similar to previous models in the disease dynamics within each age group, Arregui et al.[1] provide more detail on contact patterns between age groups and the interaction between ageing and transmission dynamics. We provide here a brief summary of the model's key aspects, including natural history of the disease, coupling between aging and disease, and uncertainty propagation. For a more in-depth description, we refer to the original publication[1].

#### *M.tb.* transmission dynamics within age strata.

The model has an age-based structure consisting of 15 age groups, with 14 groups covering a span of 5 years up to 70 years, and the last group including individuals older than 70 years. In each age group, there are unexposed individuals (susceptible), two latency paths to disease (fast and slow), and six types of disease based on their aetiology: nonpulmonary, pulmonary (smear-positive), and pulmonary (smear-negative), with or without treatment.

The model also includes WHO data schemes for treatment outcomes, such as treatment completion, default, failure, and death. The model is compartmental, simulating the natural history of TB through 19 possible reservoirs that depends upon the host's disease status. Supplementary Figure 1 illustrates the entire model. The model includes various transitions between compartments of the TB natural history, which are summarized as follows:

- Infection processes: after contact with an infectious individual, susceptible individuals ($S$) get infected, entering either the fast ($L_F$), or slow latency states ($L_S$).
- Re-infection processes: individuals in the slow latency reservoir can get re-infected, a fraction of which will develop TB fast after re-infection. This is modeled as a transition from $L_S$ to $L_F$.
- Development of active TB: infected individuals (either $L_F$ or $L_S$) may develop initially undiagnosed -and thus untreated- TB ($D$ states).
- TB diagnosis: with some delay after the disease onset, TB gets diagnosed and treatment starts (transition from $D$ states to $T$ ones)

- Treatment outcomes: (transitions from $T$ states to $R$ ones) different possible outcomes are possible: –either success or failure/default- (failure of treatment moves individuals to F)
- Disease relapse: (transitions back from $T$ to $R$)
- Death: active TB patients, either diagnosed or not, are assigned a TB-specific mortality rate.

Arregui et al.'s work[1] provides explicit details about the mathematical parameterization of the model transitions, which can be found in the supplementary appendix, while the next section describes the ODE system governing the evolution in each compartment. Altogether, they define the evolution of individuals over time, ultimately yielding global estimates of TB burden when aggregated across age strata.

Within this framework, infections occur when susceptible individuals come into contact with infectious ones. If $S(a,t)$ represents the number of susceptible individuals in the age group $a$ at time $t$, the number of new infections can be determined by multiplying $S(a,t)$ by the force of infection experienced by that sub-population, denoted as $\lambda(a,t)$. This force of infection represents the fraction of susceptible individuals who become infected per year and is proportional to the sum:

$$\sum_{a'} \xi_c(a,a',t)\Upsilon(a',t) \tag{1}$$

where $\Upsilon(a',t)$ is the density of all the infectious individuals within age-group $a'$ at time step $t$, weighted by their relative infectiousness; and $\xi_c(a,a',t)$ represents the relative contact frequency that an individual of age $a$ has with individuals of age $a'$ at time $t$, with respect to the overall average of contacts that an individual has per unit time with anyone else. For the computation of the contact matrices we capitalized on contact survey studies. In the case of China we use the survey conducted there[2] for getting a matrix with its CI that is usable in China, and that is bradly representative of the contact structure in the Asian region. The matrix granularity is adapted to match 15 age groups, as described in[1,3]. The confidence intervals of each field in the matrices is given in the study and in inherited. Importantly, we also take into account that, as the demographic structure of the population changes, the contact patterns change too[3], being this the origin of the dependence over time found in matrices.

#### Ordinary differential equations system

Supplementary Figure 1 displays the TB natural history compartments used in the model, representing the different states of the compartmental model. The population's progression through each compartment is captured by an ODE, reflecting the transitions that bring in new individuals or remove them. The evolution of the model's various dynamic states is described by the following system of differential equations:

$$\begin{aligned}\dot{S}(a,t) &= -\lambda(a,t)S(a,t) - ((1-\delta(a-14))S(a,t) - (1-\delta(a))S(a-1,t))/\tau \\ &+ \delta(a)(1-m_c m_d(t))\Delta_N(a,t) + (1-\delta(a))\Delta_N(a,t)S(a,t)/N(a,t)\end{aligned} \tag{2}$$

$$\begin{aligned}\dot{L}_s(a,t) &= (1-p(a))\lambda(a,t)S(a,t) - p(a)q\lambda(a,t)L_s(a,t) - \omega_s L_s(a,t) + \delta(a)m_c m_d(t)(1-p(0))\Delta_N(a,t) \\ &- ((1-\delta(a-14))L_s(a,t) - (1-\delta(a))L_s(a-1,t))/\tau + (1-\delta(a))\Delta_N(a,t)L_s(a,t)/N(a,t)\end{aligned} \tag{3}$$

$$\begin{aligned}\dot{L}_f(a,t) &= p(a)\lambda(a,t)S(a,t) - \omega_f L_f(a,t) + p(a)q\lambda(a,t)(L_s(a,t) + R_{p+N}(a,t) + R_{p-N}(a,t) + R_{npN}(a,t)) \\ &+ p(a)q\lambda(a,t)(R_{p+S}(a,t) + R_{p-S}(a,t) + R_{npS}(a,t) + R_{p+D}(a,t) + R_{p-D}(a,t) + R_{npD}(a,t)) \\ &- ((1-\delta(a-14))L_f(a,t) - (1-\delta(a))L_f(a-1,t))/\tau + \delta(a)m_c m_d(t)p(0)\Delta_N(a,t) \\ &+ (1-\delta(a))\Delta_N(a,t)L_f(a,t)/N(a,t)\end{aligned} \tag{4}$$

$$\begin{aligned}\dot{D}_{p+}(a,t) &= \omega_f \rho_{p+}(a)L_f(a,t) + \omega_s \rho_{p+}(a)L_s(a,t) - \mu_{p+}D_{p+}(a,t) - d(t)D_{p+}(a,t) \\ &- \nu D_{p+}(a,t) + r_N R_{p+N}(a,t) + r_S R_{p+S}(a,t) + r_D R_{p+D}(a,t) + \theta D_{p-}(a,t) \\ &- ((1-\delta(a-14))D_{p+}(a,t) - (1-\delta(a))D_{p+}(a-1,t))/\tau + (1-\delta(a))\Delta_N(a,t)D_{p+}(a,t)/N(a,t)\end{aligned} \tag{5}$$

$$\begin{aligned}\dot{D}_{p-}(a,t) &= \omega_f(1-\rho_{p+}(a)-\rho_{np}(a))L_f(a,t)+\omega_s(1-\rho_{p+}(a)-\rho_{np}(a))L_s(a,t)-\mu_{p-}D_{p-}(a,t)\\ &- \eta d(t)D_{p-}(a,t)-\nu D_{p-}(a,t)+r_NR_{p-N}(a,t)+r_SR_{p-S}(a,t)+r_DR_{p-D}(a,t)-\theta D_{p-}(a,t)\\ &- ((1-\delta(a-14))D_{p-}(a,t)-(1-\delta(a))D_{p-}(a-1,t))/\tau+(1-\delta(a))\Delta_N(a,t)D_{p-}(a,t)/N(a,t)\end{aligned} \tag{6}$$

$$\begin{aligned}\dot{D}_{np}(a,t) &= \omega_f\rho_{np}(a)L_f(a,t)+\omega_s\rho_{np}(a)L_s(a,t)-\mu_{np}D_{np}(a,t)-\eta d(t)D_{np}(a,t)\\ &- \nu D_{np}(a,t)+r_NR_{npN}(a,t)+r_SR_{npS}(a,t)+r_DR_{npD}(a,t)\\ &- ((1-\delta(a-14))D_{np}(a,t)-(1-\delta(a))D_{np}(a-1,t))/\tau+(1-\delta(a))\Delta_N(a,t)D_{np}(a,t)/N(a,t)\end{aligned} \tag{7}$$

$$\begin{aligned}\dot{T}_{p+}(a,t) &= d(t)D_{p+}(a,t)-\Psi T_{p+}(a,t)+\theta T_{p-}(a,t)\\ &- ((1-\delta(a-14))T_{p+}(a,t)-(1-\delta(a))T_{p+}(a-1,t))/\tau+(1-\delta(a))\Delta_N(a,t)T_{p+}(a,t)/N(a,t)\end{aligned} \tag{8}$$

$$\begin{aligned}\dot{T}_{p-}(a,t) &= \eta d(t)D_{p-}(a,t)-\Psi T_{p-}(a,t)-\theta T_{p-}(a,t)\\ &- ((1-\delta(a-14))T_{p-}(a,t)-(1-\delta(a))T_{p-}(a-1,t))/\tau+(1-\delta(a))\Delta_N(a,t)T_{p-}(a,t)/N(a,t)\end{aligned} \tag{9}$$

$$\begin{aligned}\dot{T}_{np}(a,t) &= \eta d(t)D_{np}(a,t)-\Psi T_{np}(a,t)\\ &- ((1-\delta(a-14))T_{np}(a,t)-(1-\delta(a))T_{np}(a-1,t))/\tau+(1-\delta(a))\Delta_N(a,t)T_{np}(a,t)/N(a,t)\end{aligned} \tag{10}$$

$$\begin{aligned}\dot{F}(a,t) &= \Psi f_F^{p+}T_{p+}(a,t)+\Psi f_F^{p-}(T_{p-}(a,t)+T_{np}(a,t))-\mu_{p+}F(a,t)\\ &- ((1-\delta(a-14))F(a,t)-(1-\delta(a))F(a-1,t))/\tau+(1-\delta(a))\Delta_N(a,t)F(a,t)/N(a,t)\end{aligned} \tag{11}$$

$$\begin{aligned}\dot{R}_{p+N}(a,t) &= \nu D_{p+}(a,t)-r_NR_{p+N}(a,t)-p(a)q\lambda(a,t)R_{p+N}(a,t)\\ &- ((1-\delta(a-14))R_{p+N}(a,t)-(1-\delta(a))R_{p+N}(a-1,t))/\tau+(1-\delta(a))\Delta_N(a,t)R_{p+N}(a,t)/N(a,t)\end{aligned} \tag{12}$$

$$\begin{aligned}\dot{R}_{p-N}(a,t) &= \nu D_{p-}(a,t)-r_NR_{p-N}(a,t)-p(a)q\lambda(a,t)R_{p-N}(a,t)\\ &- ((1-\delta(a-14))R_{p-N}(a,t)-(1-\delta(a))R_{p-N}(a-1,t))/\tau+(1-\delta(a))\Delta_N(a,t)R_{p-N}(a,t)/N(a,t)\end{aligned} \tag{13}$$

$$\begin{aligned}\dot{R}_{npN}(a,t) &= \nu D_{np}(a,t)-r_NR_{npN}(a,t)-p(a)q\lambda(a,t)R_{npN}(a,t)\\ &- ((1-\delta(a-14))R_{npN}(a,t)-(1-\delta(a))R_{npN}(a-1,t))/\tau+(1-\delta(a))\Delta_N(a,t)R_{npN}(a,t)/N(a,t)\end{aligned} \tag{14}$$

$$\begin{aligned}\dot{R}_{p+S}(a,t) &= \Psi(1-f_D^{p+}-f_F^{p+}-f_\mu^{p+})T_{p+}(a,t)-r_SR_{p+S}(a,t)-p(a)q\lambda(a,t)R_{p+S}(a,t)\\ &- ((1-\delta(a-14))R_{p+S}(a,t)-(1-\delta(a))R_{p+S}(a-1,t))/\tau+(1-\delta(a))\Delta_N(a,t)R_{p+S}(a,t)/N(a,t)\end{aligned} \tag{15}$$

$$\begin{aligned}\dot{R}_{p-S}(a,t) &= \Psi(1-f_D^{p-}-f_F^{p-}-f_\mu^{p-})T_{p-}(a,t)-r_SR_{p-S}(a,t)-p(a)q\lambda(a,t)R_{p-S}(a,t)\\ &- ((1-\delta(a-14))R_{p-S}(a,t)-(1-\delta(a))R_{p-S}(a-1,t))/\tau+(1-\delta(a))\Delta_N(a,t)R_{p-S}(a,t)/N(a,t)\end{aligned} \tag{16}$$

$$\begin{aligned}\dot{R}_{npS}(a,t) &= \Psi(1-f_D^{p-}-f_F^{p-}-f_\mu^{p-})T_{np}(a,t)-r_SR_{npS}(a,t)-p(a)q\lambda(a,t)R_{npS}(a,t)\\ &- ((1-\delta(a-14))R_{npS}(a,t)-(1-\delta(a))R_{npS}(a-1,t))/\tau+(1-\delta(a))\Delta_N(a,t)R_{npS}(a,t)/N(a,t)\end{aligned} \tag{17}$$

$$\begin{aligned}\dot{R}_{p+D}(a,t) &= \Psi f_D^{p+}T_{p+}(a,t)-r_DR_{p+D}(a,t)-p(a)q\lambda(a,t)R_{p+D}(a,t)\\ &- ((1-\delta(a-14))R_{p+D}(a,t)-(1-\delta(a))R_{p+D}(a-1,t))/\tau+(1-\delta(a))\Delta_N(a,t)R_{p+D}(a,t)/N(a,t)\end{aligned} \tag{18}$$

$$\begin{aligned}\dot{R}_{p-D}(a,t) &= \Psi f_D^{p-}T_{p-}(a,t)-r_DR_{p-D}(a,t)-p(a)q\lambda(a,t)R_{p-D}(a,t)\\ &- ((1-\delta(a-14))R_{p-D}(a,t)-(1-\delta(a))R_{p-D}(a-1,t))/\tau+(1-\delta(a))\Delta_N(a,t)R_{p-D}(a,t)/N(a,t)\end{aligned} \tag{19}$$

$$\begin{aligned}\dot{R}_{npD}(a,t) &= \Psi f_D^{p-}T_{np}(a,t)-r_DR_{npD}(a,t)-p(a)q\lambda(a,t)R_{npD}(a,t)\\ &- ((1-\delta(a-14))R_{npD}(a,t)-(1-\delta(a))R_{npD}(a-1,t))/\tau+(1-\delta(a))\Delta_N(a,t)R_{npD}(a,t)/N(a,t)\end{aligned} \tag{20}$$

where $\delta(a)$ stands for the Dirac delta function ($\delta(x = 0) = 1$ and $\delta(x \neq 0) = 0$). There are three time-dependent quantities: the force of infection $\lambda(a, t)$, the diagnosis rate $d(t)$, and the correction terms $\Delta_N(a, t)$, accounting for demographic changes in the population due to factors outside of TB and aging.

For a comprehensive description of the ODE system and its parameters, please see the supplementary materials of the original source[1]. In summary, the parameters used in this study represent probabilities or rates reported in the literature that reflect the progression of individuals through the model's various states. For example, some parameters represent the likelihood of developing a certain type of TB or the rate of progression from slow or fast latency to active TB.

#### Literature-based epidemiological parameters

Table I shows a summary of the model parameters used in the ODE system governing the evolution of individuals within the TB natural history. These parameters represent probabilities or rates that determine how individuals transition between different compartments and the population evolves over time. The parameter values are based on the literature. The table provides a brief description of each parameter and its assigned value.

| **Meaning** | **Parameter** | **Value** | **C.I.** |
|---|---|---|---|
| Probability of fast progression | $p(a)$ | $(a = 0)$ 0.187 | (0.1474,0.2333) |
| | | $(a = 1)$ 0.0225 | (0.0200,0.0250) |
| | | $(a > 1)$ 0.15 | (0.10,0.20) |
| Rate of fast progression ($y^{-1}$) | $\omega_f$ | 0.900 | (0.765,1.035) |
| Rate of slow progression ($y^{-1}$) | $\omega_s$ | $7.500\times10^{-4}$ | $(6.375,8.625)\times10^{-4}$ |
| Probability of developing pulmonary smear-positive disease | $\rho_{p+}(a)$ | $(a < 3)$ 0.100 | (0.085,0.115) |
| | | $(a \geq 3)$ 0.500 | (0.425,0.575) |
| Probability of developing non-pulmonary disease | $\rho_{np}(a)$ | $(a < 3)$ 0.250 | (0.2125,0.2875) |
| | | $(a \geq 3)$ 0.100 | (0.085,0.115) |
| Mortality rate by pulmonary smear positive TB ($y^{-1}$) | $\mu_{p+}$ | 0.250 | (0.213,0.288) |
| Mortality rate by pulmonary smear negative TB ($y^{-1}$) | $\mu_{p-}$ | 0.100 | (0.085,0.115) |
| Mortality rate by non-pulmonary TB ($y^{-1}$) | $\mu_{np}$ | 0.100 | (0.085,0.115) |
| Reduction of infection risk for previously infected individuals | q | 0.650 | (0.553,748) |
| Treatment completion rate ($y^{-1}$) | $\Psi$ | 2.00 | (1.70,2.30) |
| Smear progression rate ($y^{-1}$) | $\theta$ | 0.015 | (0.007,0.020) |
| Relapse rate for individuals who successfully completed treatment ($y^{-1}$) | $r_S$ | $9.392\times10^{-4}$ | $(6.364,12.450)\times10^{-4}$ |
| Relapse rate for individuals who defaulted treatment ($y^{-1}$) | $r_D$ | $3.774\times10^{-3}$ | $(1.354,8.620)\times10^{-3}$ |
| Relapse rate for naturally recovered individuals ($y^{-1}$) | $r_N$ | 0.030 | (0.020,0.040) |
| Natural recovery rate ($y^{-1}$) | $\nu$ | 0.100 | (0.085,0.115) |
| Infectiousness reduction coefficient of $D_{p-}$ with respect to $D_{p+}$ | $\phi_{p-}$ | 0.250 | (0.213,0.288) |
| Infectiousness reduction coefficient of $R_{p+D}$ with respect to $D_{p+}$ | $\phi_D$ | 0.500 | (0.250,0.750) |
| Proportion of mothers that infect their newborn children | $m_c$ | 0.15 | (0.10,0.20) |
| Diagnosis rate reduction of $D_{p-}$ and $D_{np}$ with respect to $D_{p+}$ | $\eta$ | Ethiopia, Nigeria: 0.843 | (0.664,1.022) |
| | | India, Indonesia: 0.797 | (0.628,0.966) |

TABLE I. Bibliography-based epidemiological parameters used in this study.

**Treatment outcomes probabilities**

The probabilities of individuals to end their treatment according the four categories defined by the WHO (success, default, failure or death), defined as:

- $(f_D^{p+}, f_F^{p+}, f_\mu^{p+})$: fraction of default, failure and death outcomes for smear positive pulmonary TB.

- $(f_D^{p-}, f_F^{p-}, f_\mu^{p-})$: fraction of default, failure and death outcomes for smear negative pulmonary and non pulmonary TB.

have been obtained from the WHO Treatment Outcomes database for each country, and their values in China are presented in table II:

| **Parameter** | **Value in China** |
|---|---|
| $f_D^{p+}$ (%) | 0.59 (0.58,0.60) |
| $f_F^{p+}$ (%) | 0.74 (0.73,0.75) |
| $f_\mu^{p+}$ (%) | 1.35 (1.34,1.36) |
| $f_D^{p-}$ (%) | 1.03 (1.02,1.04) |
| $f_F^{p-}$ (%) | 0.197 (0.192,0.203) |
| $f_\mu^{p-}$ (%) | 1.02 (1.01,1.03) |

TABLE II. Values of the treatment outcomes probabilities in China

**Population dynamics across age strata: individuals' ageing and demographic evolution.**

The model presented in this work characterizes disease dynamics across all age groups and incorporates age-dependent parameters. It not only tracks the evolution of sub-populations associated with disease states, but also accounts for aging dynamics, in which individuals move between different age strata as they age. This phenomenon is illustrated in Supplementary Figure 2 through the transitions between age strata and the changing shape of the demographic pyramid over time.

Two additional ingredients that are key to describing the evolution of the population and the aging are then included in our model. First, empirical data and forecasts are used to model past and future fertility levels. Second, continuous correction terms, $\Delta_N(a,t)$, are included in the model to adjust the population within age strata $a$ at time $t$ during the simulation.

To ensure the model captures the demographic prospects, the correction terms $\Delta_N(a,t)$ are dynamically calculated to match the demographic forecasts in the UN population division database. These terms are proportionally distributed among disease states based on their size, and represent changes in the population unrelated to TB dynamics (such as mortality and migration).

Supplementary Figure 2 illustrates the evolution of demographic pyramids, where individuals transition between age strata as they age, and the whole population is updated according to UN population division data, where the correction terms $\Delta_N(a,t)$ are used for this purpose.

Further details are provided in the supplementary materials of the original paper[1].

**Interaction of vaccines and the natural history of TB**

In Figure 3 we schematize the effects of the vaccines in halting transmission between states of the model. Only those individuals that are protected, among all vaccinated individuals, will see a modified version of the natural history, with the remaining vaccinated but unprotected individuals, and non-vaccinated individuals, evolving according to Figure 3A. This is due to the vaccine being modelled as all-or-nothing, where only a fraction of the vaccinated individuals, given by the efficacy, get protection from the vaccine. For those individuals, the efficacy in halting progression to disease in the vaccine mechanism is perfect. In Figure 3, panels **B, C, D, E, F** we depict the modifications that the vaccine imposes in the natural history for POD or POR vaccines acting through the mechanisms of the main text.

#### Breaking down TB cases according to its origin

In this work we estimated the distribution of TB cases across the different routes to disease that are classically described in TB. Then, we are taking into account the following cases: fast progression to primary TB upon recent infection, TB upon exogenous reinfection, TB after endogenous reactivation from LTBI, or TB recurrence, after a previous disease event. In the following lines, we summarize how each contribution is estimated to build the final results in Figure 4 of the main text.

In our model, the cases corresponding to TB after endogenous reactivation from LTBI can be estimated easily, as they map directly to the fluxes between the reservoir $L_S$, and the reservoirs $D_{p+}$, $D_{p-}$ and $D_{np}$, as showed in Supplementary Figure 1. This way, the contribution at any time $t$ to the overall pool of TB cases will be calculated as:

$$C_{\text{endogenous}}(t) = \sum_i f_{L \to D_i}(t) \tag{21}$$

Similarly, the cases corresponding to TB recurrence are also directly mappable to specific transitions between reservoirs, being those the ones between $R_x$ reservoirs and $D_i$ ones. The contribution at any time $t$ to the overall pool of TB cases will be calculated as:

$$C_{\text{recurrence}}(t) = \sum_i, x f_{R_x \to D_i}(t) \tag{22}$$

However, for primary TB and exogenous reactivation cases it is not possible to get a 1 to 1 correspondence in terms of the transitions in the model, as all individuals ends in $L_F$ reservoir as a middle step in its way to disease, effectively mixing all the individuals together, which make them indistinguishable. To solve this, we approximate the contribution to disease using the fluxes between the corresponding origin and the fast latency reservoir $L_F$, times the rate of fast progression to disease $r$, as the individuals are not expected to stay for a large amount of time in this reservoir, and they will abandon it precisely at this rate $r$.

Then, the contribution of primary TB is estimated as:

$$C_{\text{primary}}(t) = r \cdot f_{S \to L_F}(t) \tag{23}$$

whereas the contribution of exogenous reinfection can have two origins, a reinfection while in latency or a reinfection after treatment, so the final estimator is build as:

$$C_{\text{exogenous}}(t) = r \cdot \left[ f_{L_S \to L_F}(t) + \sum_x f_{R_x \to L_F}(t) \right] \tag{24}$$

Then, with each one of those contributions calculated independently, it is possible to build the distribution of cases at any time $t$.

### SUPPLEMENTARY RESULTS

#### Evolution of contact matrices using each method

In Supplementary Figure 4A, we included the results after updating the contact matrix using the pairwise correction to obtain an usable matrix in the model in the year 2010. In In Supplementary Figure 4B, we shown the projection of this same matrix to the year 2050, when the same pairwise method is used. In Supplementary Figure 4, panels C and D, we present analogous results but, instead of using the simple pairwise correction method, we compute the matrices using the intrinsic connectivity method for the years 2010 and 2050 respectively.

It is noticeable that, when evolution of demography is captured in the contact matrix using the intrinsic connectivity method, elder population tends to contact more than when using the pairwise correction, whereas adolescents tend to contact less. This could be the main driver mechanism of the inversion of the force of infection in the two age groups analyzed, which is depicted in Figure 4 of the main text.

#### Impact of vaccines with 20 years of waning

To further contextualize the findings in the main text we produce an additional analysis focused on repeating the forecast of all vaccines in all populations, but for a greater level of waning. The results, for a 20y waning, is shown in

Supplementary Figure 5. The hierarchy of impacts is the same as in the main text, and we reproduce again the fact that, if the vaccine is able to protect naive individuals, both adolescents and elders show great impacts, whereas for a vaccine protecting already exposed individuals, only elder vaccination show promising levels of IRR.

Moreover, we also recover the same behaviour as in the cases explored in the main text, namely, that for the elder population, updating the contact matrices using the intrinsic connectivity method always gives greater impacts than with the pairwise method, and the opposite happens with adolescents, whenever the vaccine is targeting transmission (either $E_p$ or $E_q$. Finally, when dealing withe naive individuals, the updating method is relevant enough to substantially change the hierarchy of impacts between adolescents and elders.

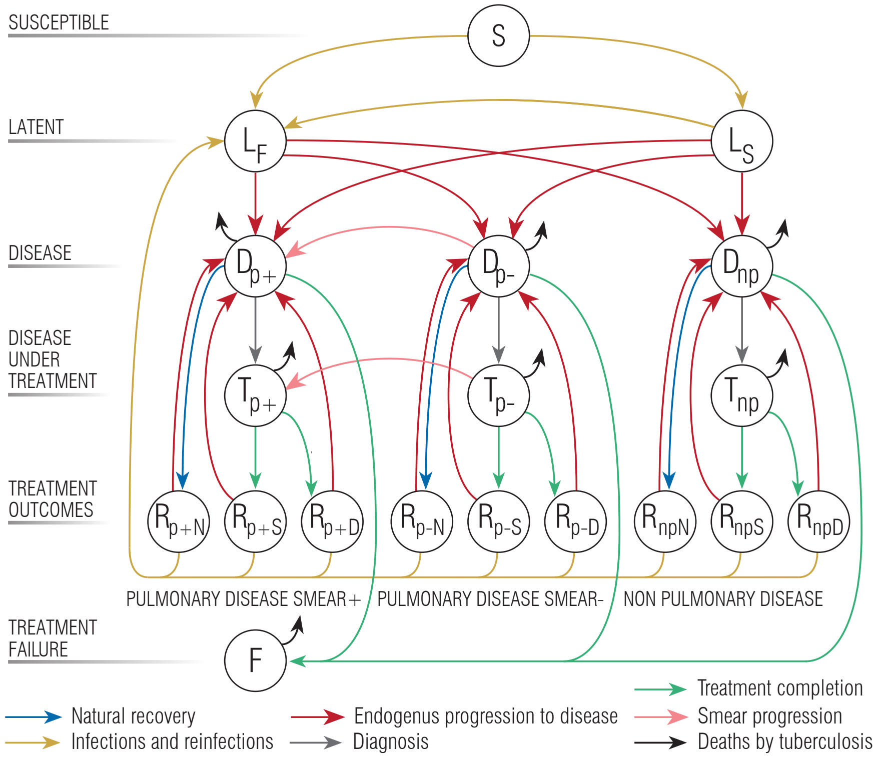


Supplementary Figure 1. Natural history of the disease employed in the mathematical model[1]. Individuals are classified according to their epidemiological status, which can be: S: susceptible. L: latent. D: (untreated) disease, T (treated) disease, R recovered, F: failed recovery . The types of TB considered here are: p+: Pulmonary Smear-Positive, p-: Pulmonary Smear-Negative, and np: Non-pulmonary. Finally, individuals under treatment end it for three different sub-types of recovery, which are: N: Natural, S: Successful, and D: Default (abandon of treatment).

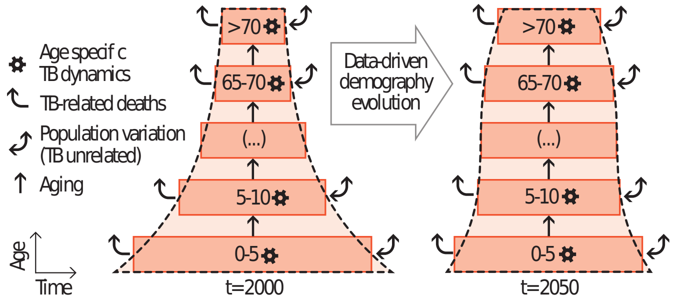


Supplementary Figure 2. Scheme of the coupling between TB dynamics and demographic evolution. The transmission model summarized in A describes the evolution of the disease in each age group, including the removal of individuals due to TB mortality (curved arrows). The evolution of the total volume of each age stratum is corrected (bidirectional arrows: TB-unrelated population variations) to make the demographic pyramid evolve according to UN prospects.

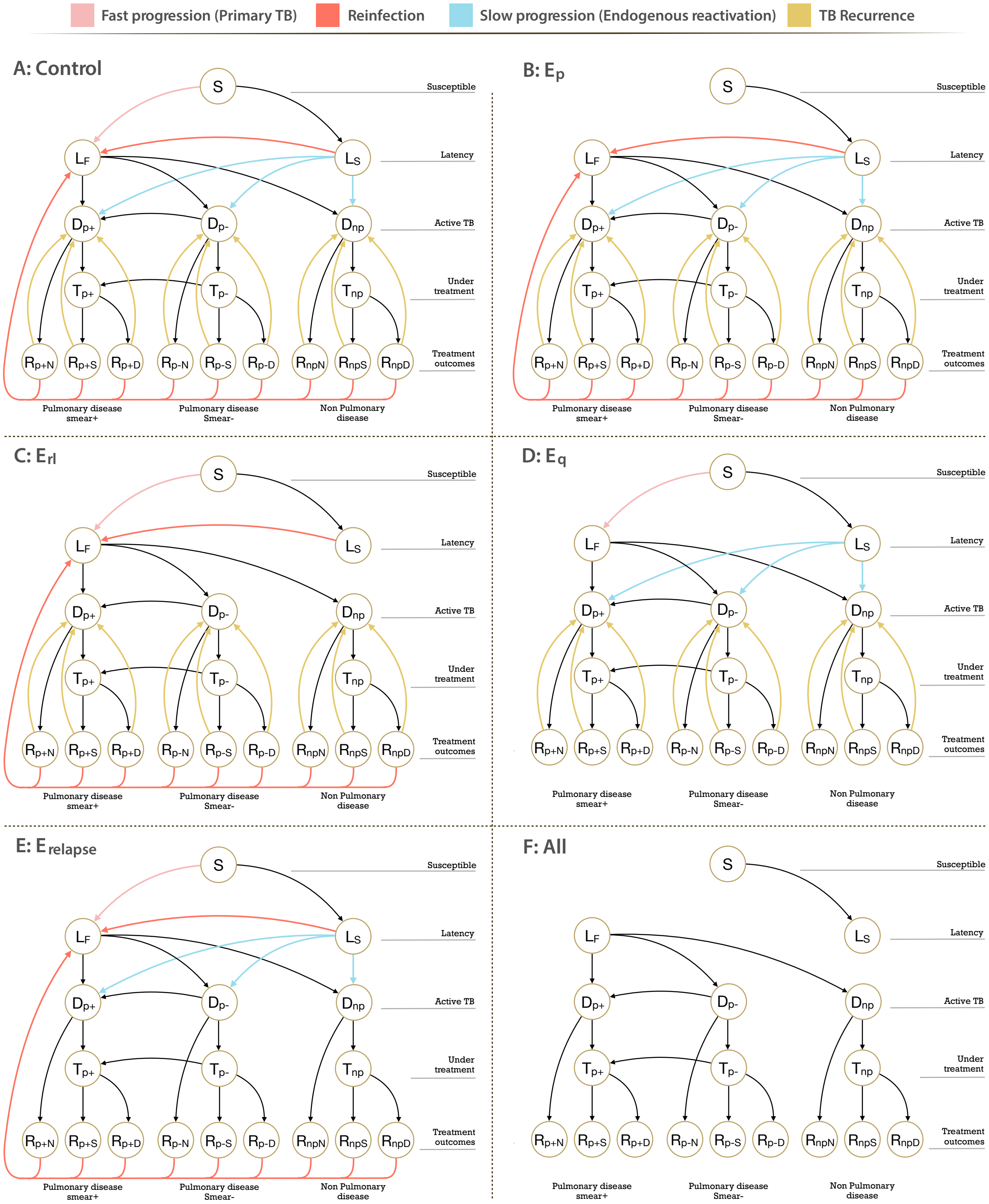


Supplementary Figure 3. Scheme of the coupling between the vaccines and TB natural history. All vaccines all all-or-nothing, and can act by halting one or more transitions between compartments. **A**. Possible transitions between compartments for unprotected individuals in the control and vaccine runs. Vaccinated and protected individuals will face a modified version of the natural history according to the mechanistic action of the vaccine. **B**. Modified natural history for an $E_p$ vaccine conferring protection against fast progression to disease upon a recent first infection event. **C**. Modified natural history for an $E_{rl}$ vaccine conferring protection against endogenous reactivation of LTBI individuals. **D**. Modified natural history for an $E_q$ vaccine conferring protection against reinfections. This vaccine prevents secondary events of infection in subjects who had been previously infected. **E**. Modified natural history for an $E_{\text{relapse}}$ vaccine conferring protection against TB recurrence. This prevents endogenous reactivation in individuals who had a past episode of active TB. **F**. Modified natural history for a vaccine conferring protection in all previous mechanisms at the same time.

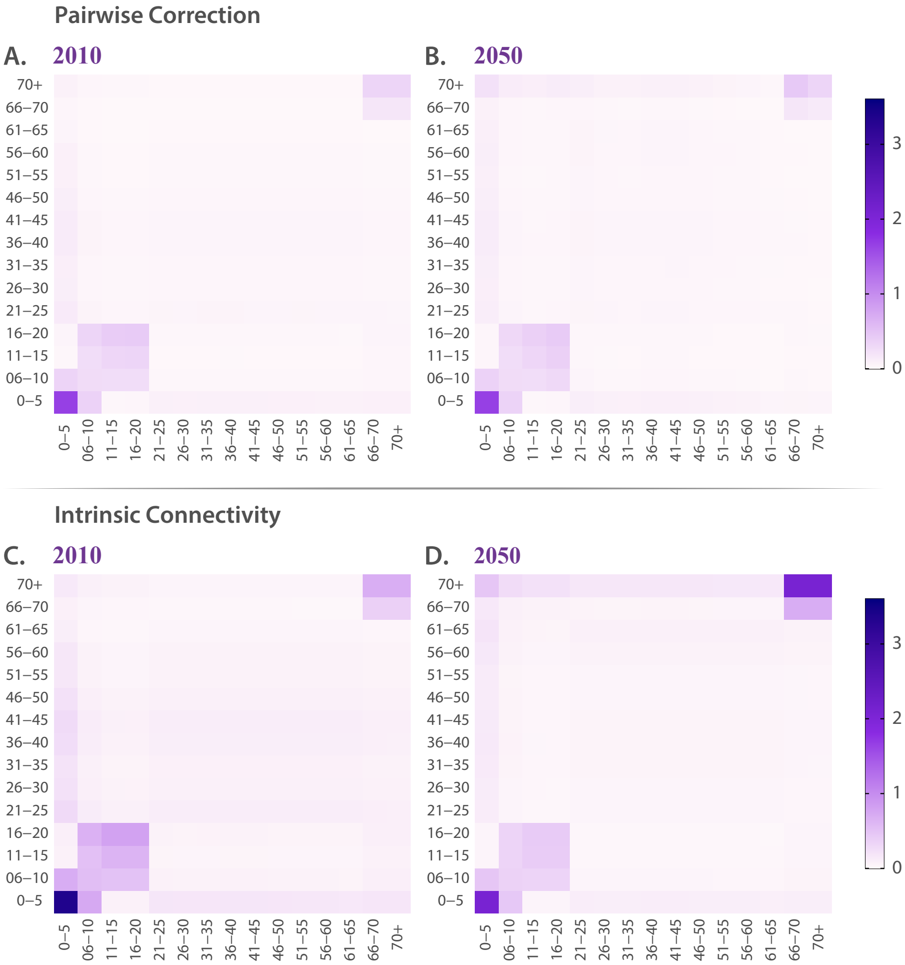


Supplementary Figure 4. Contact matrices in 2010 and projection in 2050 under selected contact update schemes (Pairwise correction and Intrinsic Connectivity). All cells capture the mean number of contacts that individuals of age $i$ have with individuals of age $j$, during a day, after the correction of the selected method is applied. When evolution of demography is captured in the contact matrix using the intrinsic connectivity method, elder population tends to contact more than when using the pairwise correction, whereas adolescents tend to contact less.

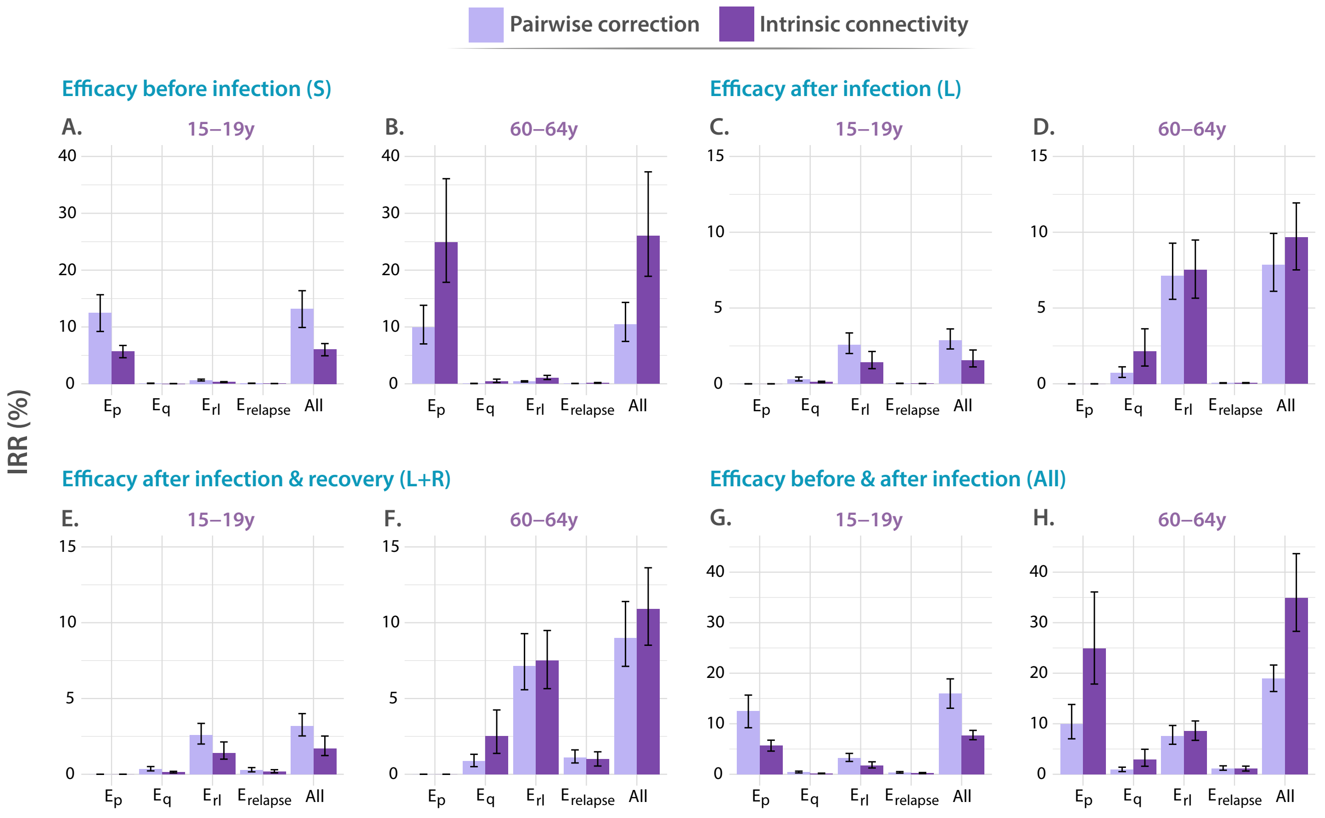


Supplementary Figure 5. Comparison between the IRR achieved when vaccinating individuals of the 15-19 or 60-64 strata. In all panels, tested vaccines act in different parts of the natural history of TB, halting progression to disease in one or more of the possible routes to disease: $E_p$: protection against primary TB, $E_q$ protection against reinfection, $E_{r_l}$: protection against endogenous progression to TB after LTBI, $E_{relapse}$: prevention of recurrence. In each case, the impact of each vaccine is evaluated for a waning level of 10 years. We analyze independently the impact of vaccines whose protective effects unfold when applied to individuals belonging to different compartments of the natural history, **A,B**. Susceptible subjects (efficacy observed before infection). **C,D**. Latently infected individuals (efficacy observed after infection). **E,F**. Latently infected and recovered individuals. **G,H**. Entire population. In each case, the impact of each vaccine is evaluated for a waning level of 10 years. In all panels, bars represent median values for the IRR measured in 2050, associated with the introduction of the vaccine in 2025. Errorbars capture 95% confidence intervals from a set of $N = 500$ model outcomes in each case.
